# Effect of physical inactivity and tobacco use on mortality and morbidity in revascularized patients with peripheral arterial disease: A nationwide cohort study

**DOI:** 10.1101/2021.05.12.21257135

**Authors:** Seungwoo Cha, Sherry L Grace, Kyungdo Han, Bongseong Kim, Nam-Jong Paik, Won-Seok Kim

## Abstract

**Importance:** Physical activity (PA) and tobacco use are key health behaviours in patients with peripheral arterial disease (PAD). Limited studies are available on effects of those behaviours in PAD after revascularization, including Asian countries where tobacco use is high.

**Objective:** To investigate the effects of PA and tobacco use on adverse clinical outcomes in patients with PAD after revascularization.

**Design:** Retrospective cohort study

**Setting:** Population-based study using the Korean National Health Insurance Service (NHIS) database

**Participants:** Patients who had received revascularization for PAD between 2010-2015 were included. They were categorized as active or inactive based on the number of days per week they engaged in PA and as current or non-tobacco users (self-report).

**Exposures:** PA and tobacco use.

**Main outcomes:** The primary outcome was all-cause mortality. Secondary outcomes included major adverse outcome (a composite of all-cause mortality, myocardial infarction, and stroke) and major adverse limb event (MALE, a composite of amputation and recurrent revascularization).

**Results:** The cohort comprised 8324 patients (mean age, 64.7 years; 76.9% male). Among them, 32.7% were inactive and 26.4% were tobacco users. Active patients had significantly better outcomes than inactive patients [all-cause mortality adjusted hazard ratio (adjHR) = 0.766 (0.685 – 0.855), major adverse outcome adjHR = 0.795 (0.719 – 0.878), MALE adjHR = 0.858 (0.773 – 0.953)]. Tobacco users had significantly poorer outcomes than non-users [all-cause mortality adjHR = 1.279 (1.124 – 1.456), major adverse outcome adjHR = 1.263 (1.124 – 1.418), MALE adjHR = 1.291 (1.143 – 1.458)].

**Conclusions and Relevance:** Even after receiving revascularization for PAD, a sizable proportion of patients were inactive and used tobacco, leading to adverse clinical outcomes. These modifiable risk factors are systematically addressed in cardiac rehabilitation; in line with current guideline recommendations, more needs to be done to ensure cardiac rehabilitation participation in patients with PAD.

## Introduction

Peripheral arterial disease (PAD) is a progressive atherosclerotic disease in arterial segments. There are more than 200 million people suffering from PAD worldwide^1^, with the highest prevalence in the Western Pacific^2^. PAD negatively affects quality of life as well as functional status, inducing pain and limitations to walking ^3,4^. Moreover, prognosis is poor, with a high incidence of cardiovascular comorbidities, such as myocardial infarction (MI) and stroke, and an annual mortality rate as high as 8.2%^5–7^.

For the management of PAD, revascularization combined with guideline-indicated pharmacotherapy, including antiplatelet and statin agents, is recommended^8,9^. With advances in technology, various endovascular procedures using stents and balloons are available, and their use is growing substantially compared to that of open bypass surgery^10,11^. In addition, exercise therapy has been shown to improve pain-free and maximum walking distance^12^; indeed, previous studies have shown that exercise therapy after endovascular procedures is superior in terms of maximum walking distance and quality of life improvement when compared to revascularization alone^13,14^. Therefore, exercise therapy is highly recommended after revascularization^15^, and cardiac rehabilitation is recommended for PAD patients^16^, although evidence about its impact on reduction of mortality rates or limb events such as recurrent revascularization or amputation is still lacking. Unfortunately, research has shown that the degree of physical activity (PA) after PAD remains low^17,18^.

Regarding other key health behaviours, tobacco use is closely associated with PAD^19^. Tobacco cessation is strongly recommended for all patients with PAD, as it leads to increased walking distance and amputation-free survival^8,9^. In a few recently published observational studies, about 30% of patients with PAD were tobacco users, and less than half of the users were able to quit successfully^20,21^. Despite the availability of effective cessation interventions, there is dearth of real-world data about tobacco use in patients with PAD, especially in Asian countries where the rates of tobacco use differ from Western countries^22^.

This study aimed to investigate the effect of PA and tobacco use in PAD patients on clinical outcomes including all-cause mortality, MI, stroke, minor and major amputation, and recurrent revascularization in a large, representative, population-based cohort in an Asian country. It was postulated that PA would have a beneficial effect, while tobacco use would have a harmful effect on adverse clinical outcomes. If demonstrated, given these risk factors are modifiable through proven approaches, efforts to address them could then lessen mortality and morbidity.

## Methods

### Data source

The Korean National Health Insurance Service (NHIS) is a single-payer organization covering the entire population. The NHIS reimburses primary and specialty care such as hospitalizations and emergency department visits in Korea. About 97% of citizens are covered by the NHIS, and the remaining 3% with the lowest income are covered by Medicaid (they are also included in the NHIS database). Information about sociodemographic characteristics (including socioeconomic status), past medical history, and mortality is collected for the NHIS^23^. The database also contains information about healthcare claims, including diagnoses as per the International Classification of Diseases, 10th revision, Clinical Modification (ICD-10-CM) codes^24^.

The NHIS provides a biennial health check-up, and participation rates reach 74.8%^25^. Through this program, height, weight, and blood pressure are measured; laboratory tests including blood cell count and chemistry are completed; and a self-administered questionnaire about medical history and health-related behaviours is collected.

This study was exempt from review by the Seoul National University Bundang Hospital Institutional Review Board (IRB number X-2001-591-901), complying with the requirements of the Declaration of Helsinki. Data from the Korean NHIS were fully anonymized for analyses, and the need for informed consent was waived.

### Study cohort

Records of patients admitted for revascularization with PAD (presence of open bypass codes O0163, O0164, O0165, O0166, O0167, O0168, O0169, O0170, O1645, O1646, O2064, O2067, O2065, O2068 and endovascular procedural codes M6597, M6605, M6613, M6620)^11^, from 1 January 2010 to 31 December 2015 were screened (**eFigure 1** in **Supplement**). Those who participated in health check-ups after revascularization within 2 years were included in the cohort for this study.

Exclusion criteria were as follows: (1) age < 40 years; (2) past medical history of MI (ICD-10-CM codes I21 and I22) and stroke (ICD-10-CM codes I60, I61, I62, I63, and I64); (3) history of minor and major limb amputation (procedural codes N0562, N0564, N0565, N0566, N0571, N0572, N0573, N0574, and N0575); and (4) occurrence of MI, stroke, recurrent revascularization, and amputation during the period from the day of revascularization therapy to the day of health check-up (**eFigure 1** in **Supplement**).

### Measures

#### PA and tobacco use

A self-administered questionnaire about leisure-time PA using a 7-day recall method was given at the time of the health check-up (**eFigure 2** in **Supplement**). The questionnaire was in Korean and consisted of three items similar to the assessment by Smith et al.^26^; the validity and reliability have been etablished^27^. PA level was classified as ‘active’ or ‘inactive’ based on the number of days per week patients engaged in PA (**eFigure 3** in **Supplement**), similar to previous research^28^.

In addition, PA-related energy expenditure (MET-min/week) was calculated by summing the multiples of frequency, intensity, and duration of PA. Light-intensity (walking), moderate-intensity, and vigorous-intensity PA thresholds used were 2.9, 4.0, and 7.0 METs, respectively^29^. Then, PA-related energy expenditure was categorized into quartiles.

Patients were categorized as current users or non-users based on responses to the questionnaire. In addition, non-users were further classified as never users or former users, and current users were classified based on degree of use (i.e., 1 to 19 cigarettes per day, or 20 or more cigarettes per day), resulting in 4 categories.

Finally, further analysis was performed with subgroups based on the combination of PA and tobacco status; inactive tobacco user, inactive non-user, active tobacco user, and active non-user.

#### Clinical outcomes

Adverse clinical outcomes after health check-up were followed in the study population until December 31, 2018. The NHIS database is linked to Statistics Korea, which records the death of any Korean citizens. Death from any cause was considered. Presence of MI and stroke was defined as admission with claims with diagnostic code I21-22 and I60-64, respectively. Major adverse outcome was defined as any of mortality, MI, and/or stroke.

Recurrent revascularization and amputation were also defined through confirmation of procedural codes, as stated previously for revascularization therapy, as well as minor and major amputation, respectively. Then, major adverse limb event (MALE) was defined as any recurrent revascularization and/or amputation.

#### Covariate data

Sociodemographic data about age, sex, income, and rural residence were extracted. Income was categorized in quartiles, based on health insurance premium; the Medicaid group was classified separately. Patients were considered to be living in a rural area if they resided in an area with a population less than 50,000 as per the Korean Local Autonomy Act.

Patients were categorized as non-drinker, mild drinker (≤30 g alcohol per day), or heavy drinker (>30 g alcohol per day)^30^, based on responses to the questionnaire. Waist circumference was categorized by the cut-off values for central obesity (90 cm for males and 80 cm for females in Asia) according to the WHO guideline^31^. Laboratory data on low-density lipoprotein and fasting plasma glucose were dichotomized as high vs. low based on PAD guidelines^8,9^.

Past medical history of end-stage renal disease, diabetes mellitus, hypertension, and dyslipidaemia was confirmed through the corresponding diagnostic codes and medication prescription history. By reviewing each patient’s ICD-10-CM diagnoses, the Charlson Comorbidity Index (CCI) was also calculated^32^. Preoperative CCI was categorized as 0, 1, or ≥ 2. Claim data about medication including antiplatelet agents (aspirin or clopidogrel), and lipid-lowering agents (such as statin, fibrate, and ezetemibe) during the year after revascularization were extracted.

### Statistical analyses

The characteristics of the study population were described using means with standard deviation, or numbers with percentages. To compare categorical and continuous variables between the ‘active’ and ‘inactive’ groups, chi-square and student’s t-tests were used, as applicable. Cox proportional hazard regression analysis was used to evaluate the effect of PA and tobacco use on adverse clinical outcomes adjusted for age, sex, procedure type, income quartile, rural residence, past medical history, CCI, medication use, length of hospital stay, days from revascularization to health check-up, and the categorized waist circumference, fasting plasma glucose, and low-density lipoprotein. Kaplan-Meier curves were also plotted in groups classified by PA and tobacco use. Statistical analyses were performed using SAS version 9.4 (SAS Institute Inc, Cary, NC, USA). A two-sided p-value < 0.05 was regarded as statistically significant.

## Results

### Cohort characteristics

A total of 8324 patients were included in the analysis (**eFigure 4** in **Supplement**). The mean age was 64.7 years, and 76.9% were male. They predominantly underwent endovascular procedures (85.0%), followed by open bypass surgery (11.9%), and combined open bypass and endovascular procedures (3.1%).

Days from revascularization to the health check-up are shown in **Table 1**. In the cohort, 2721 patients (32.7%) were inactive, and 2193 (26.3%) were tobacco users (**Table 1**). Inactive patients were significantly older, more often female, had lower income, and were more often living in a rural area than their active counterparts. They also had more end-stage renal disease, greater waist circumference, more frequent tobacco use, and longer hospital stays. Procedure type and alcohol consumption were also significantly different in the two PA groups (**Table 1**).

**Table 1.**
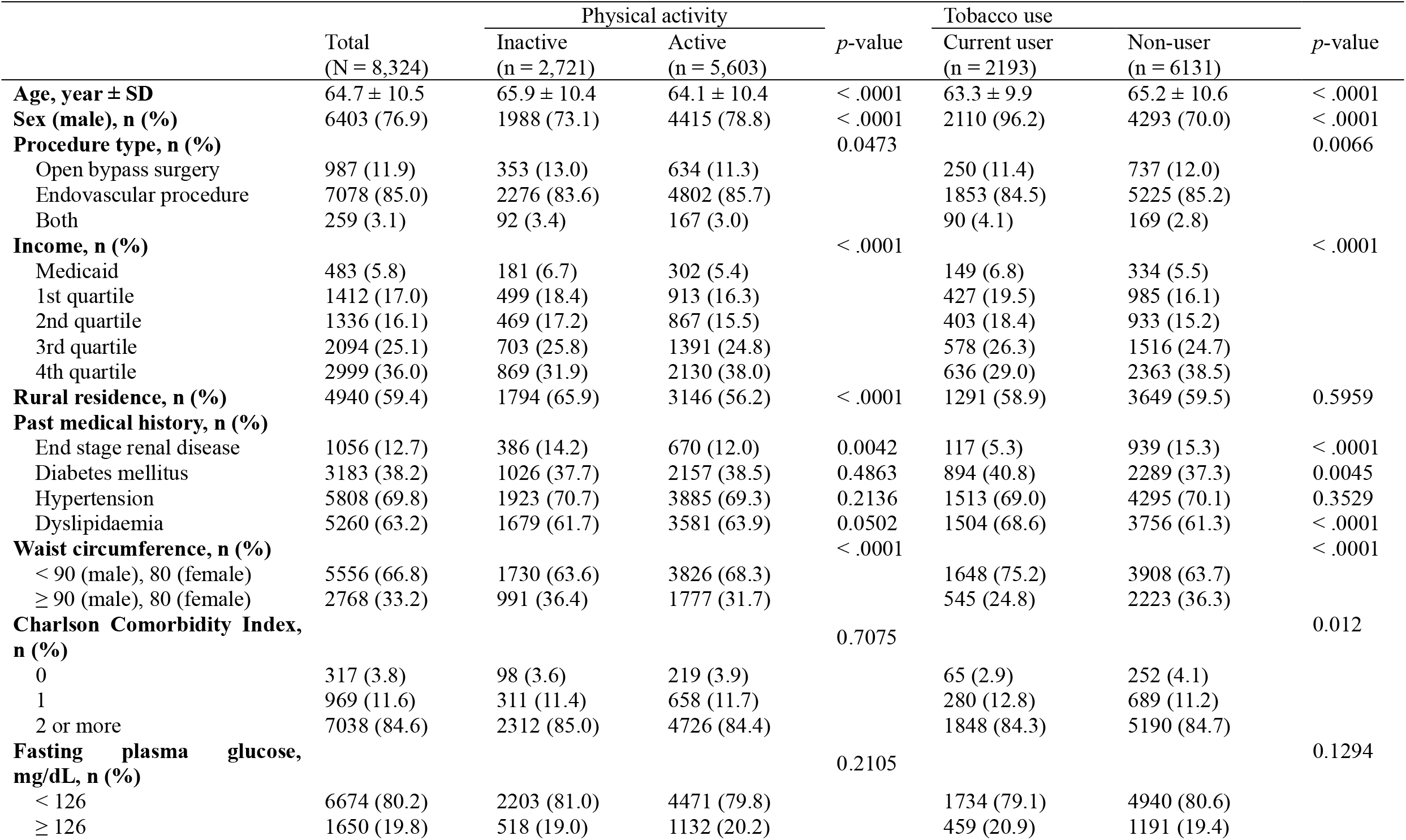
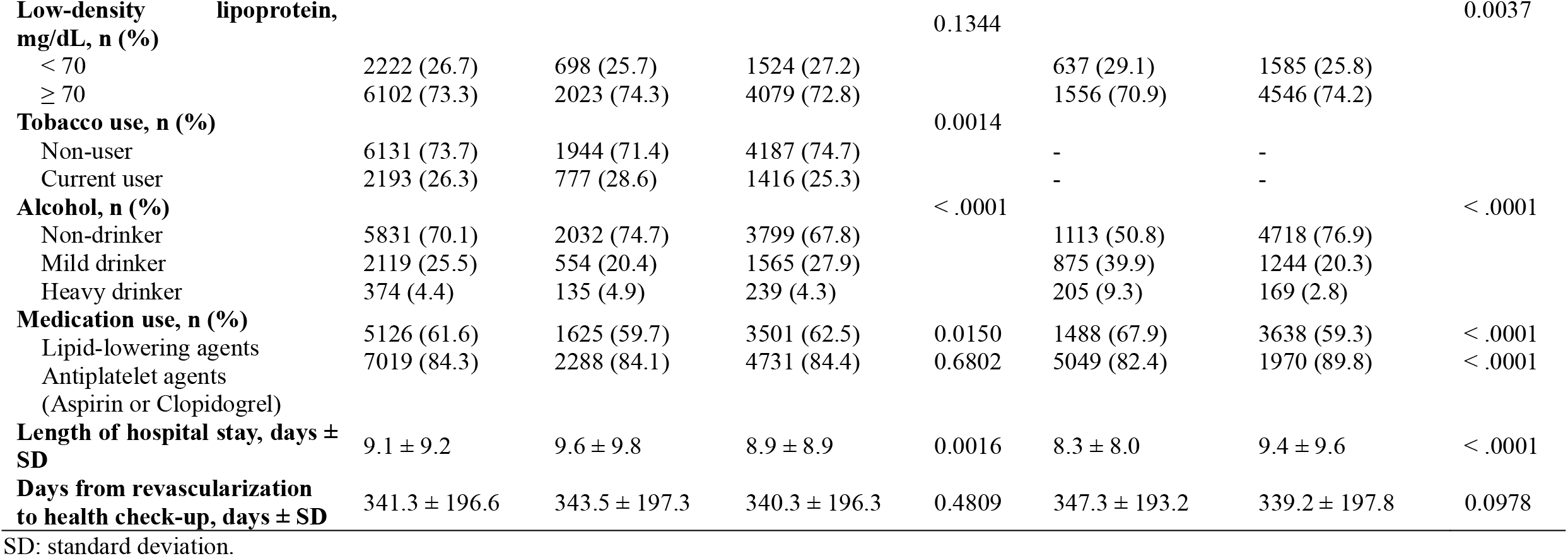
Cohort sociodemographic and clinical characteristics by physical activity and tobacco use

Tobacco users were significantly younger, were more often male, and had lower income than non-users. They also had more diabetes mellitus and dyslipidaemia, higher CCI, and higher low-density lipoprotein levels. Moreover, they had less end-stage renal disease, smaller waist circumference, and shorter hospital stays. Procedure type and alcohol consumption were also significantly different between tobacco users and non-users (**Table 1**).

### Effect of PA and tobacco use on clinical outcomes

The median and maximum follow-up periods were 4.4 years and 8.8 years, respectively. During the follow-up period, 1338 deaths (16.1%), 552 MI and/or strokes (6.7%), and 1679 recurrent revascularizations and/or amputations (20.2%) occurred.

The unadjusted Kaplan-Meier analysis showed a significant difference in clinical outcomes by PA, but not by tobacco status (**eFigure 5** in **Supplement**). In the Cox proportional hazard regression analysis, active patients showed significantly lower death [adjusted hazard ratio (adjHR) 0.766 (0.685 – 0.855)], major adverse outcome [adjHR 0.795 (0.719 – 0.878)], and MALE [adjHR 0.858 (0.773 – 0.953)], while tobacco users had significantly higher death [adjHR 1.279 (1.124 – 1.456)], major adverse outcome [adjHR 1.263 (1.124 – 1.418)], and MALE [adjHR 1.290 (1.143 – 1.458)] (**Table 2**).

**Table 2.**
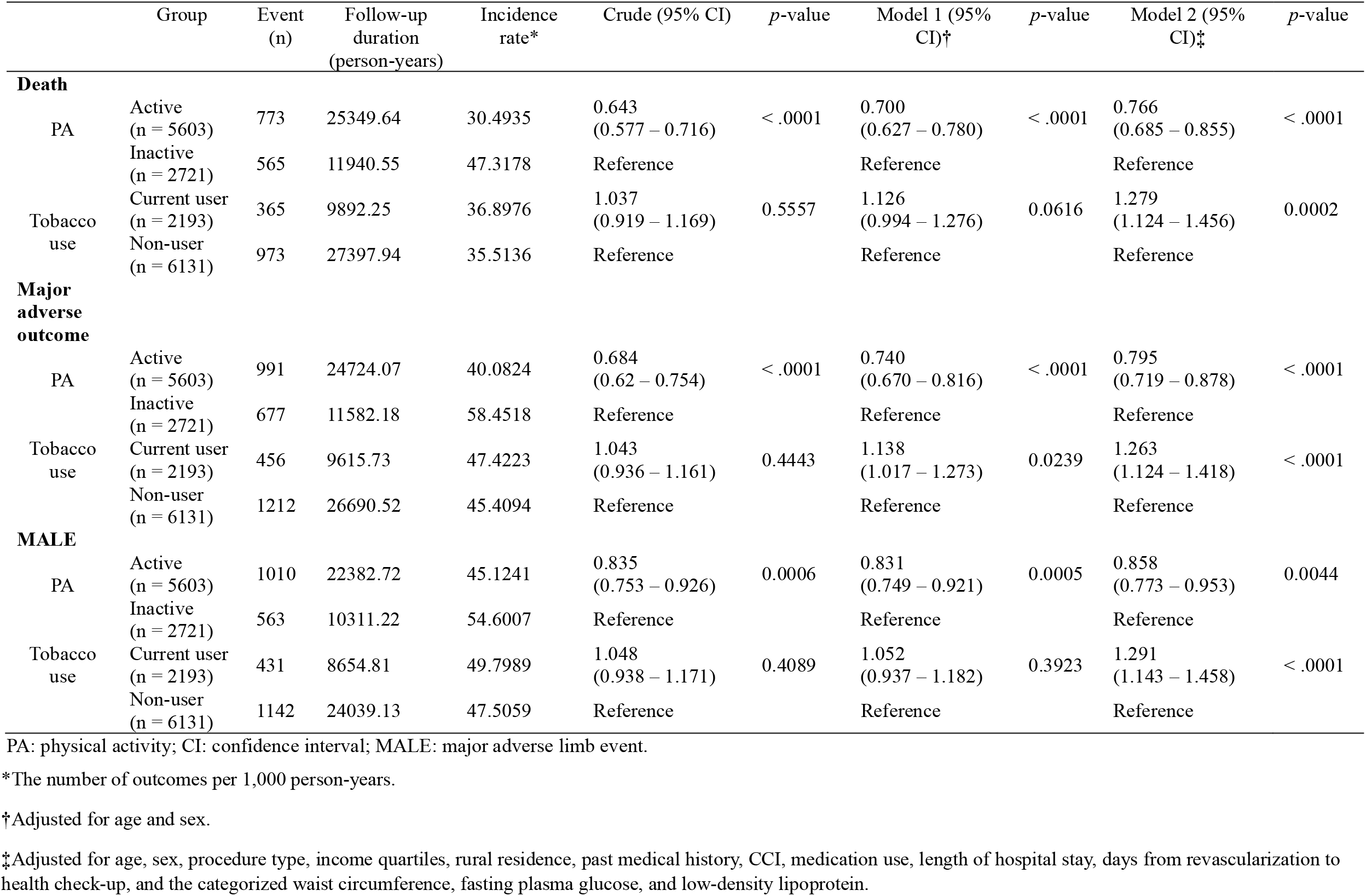
Hazard ratios for outcomes according to PA and tobacco use

When PA-related energy expenditure (MET-min/week) was examined as quartiles, higher METs were associated with significantly lower hazard of death and major adverse outcome (**Table 3**). The Kaplan-Meier analysis also revealed a significant difference in clinical outcomes (**eFigure 6** in **Supplement**). The highest MET quartile was associated with significantly lower adjHRs than the lowest quartile [death; 0.593 (0.505 – 0.696), major adverse outcome; 0.636 (0.553 – 0.732), MALE; 0.859 (0.747 – 0.987)]. The decrease in adjHR based on PA-related energy expenditure was statistically significant for death, major adverse outcome, and MALE (**Table 3**).

**Table 3.**
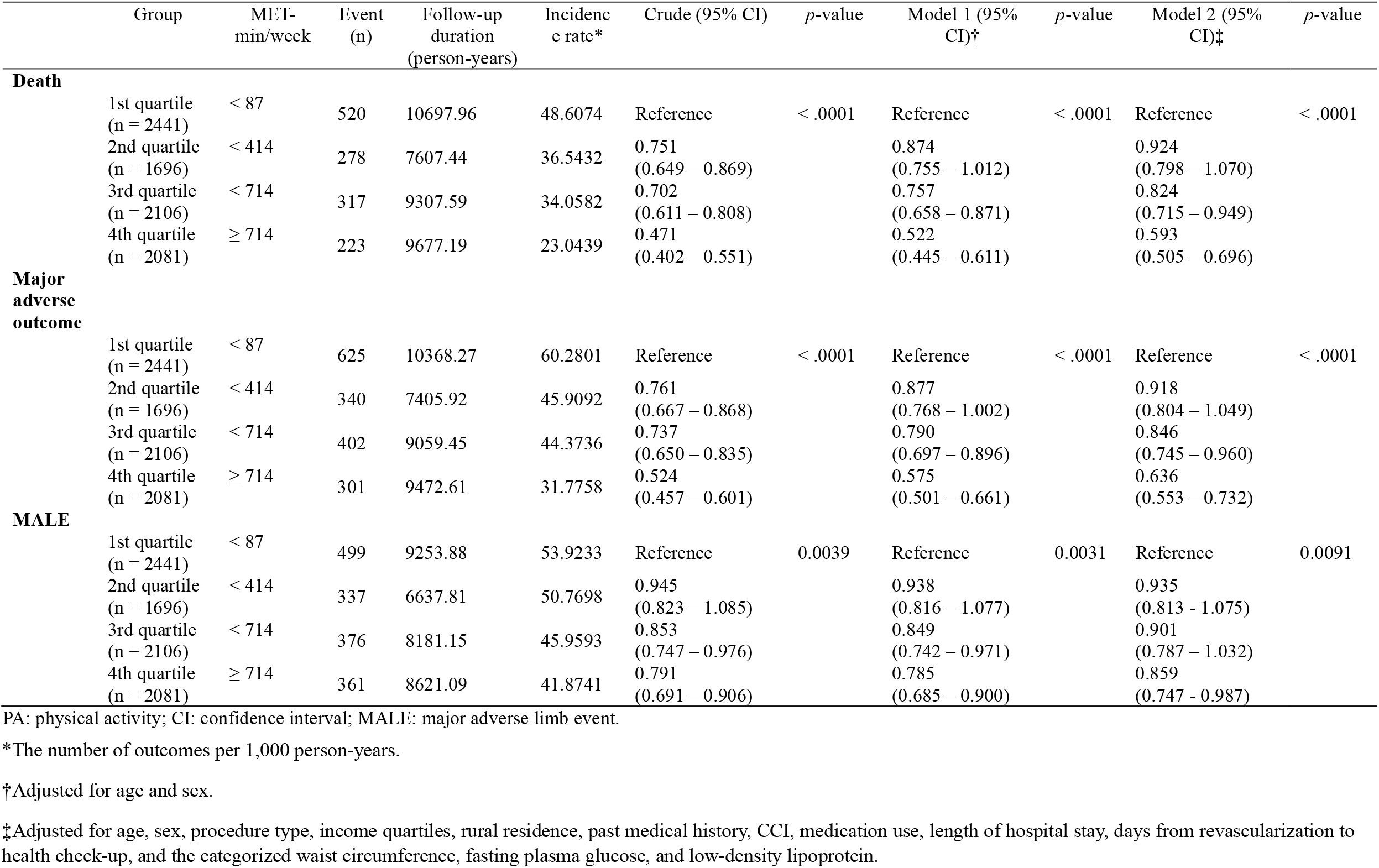
Hazard ratios for outcomes by PA-related energy expenditure (MET-min/week)

When tobacco use was further classified (never user, former user, 1 to 19 cigarettes per day, and 20 or more cigarettes per day), the latter two smoking groups showed higher adjHRs than the former two groups (**Table 4**). The group smoking 20 or more cigarettes per day showed significantly higher adjHRs than the group of never users [death; 1.210 (0.989 – 1.481), major adverse outcome; 1.226 (1.025 – 1.467), MALE; 1.354 (1.125 – 1.631)]. There was no significant difference in clinical outcomes between former and never users. The Kaplan-Meier analysis showed a significant difference in death and major adverse outcome, but not in MALE (**eFigure 7** in **Supplement**).

**Table 4.**
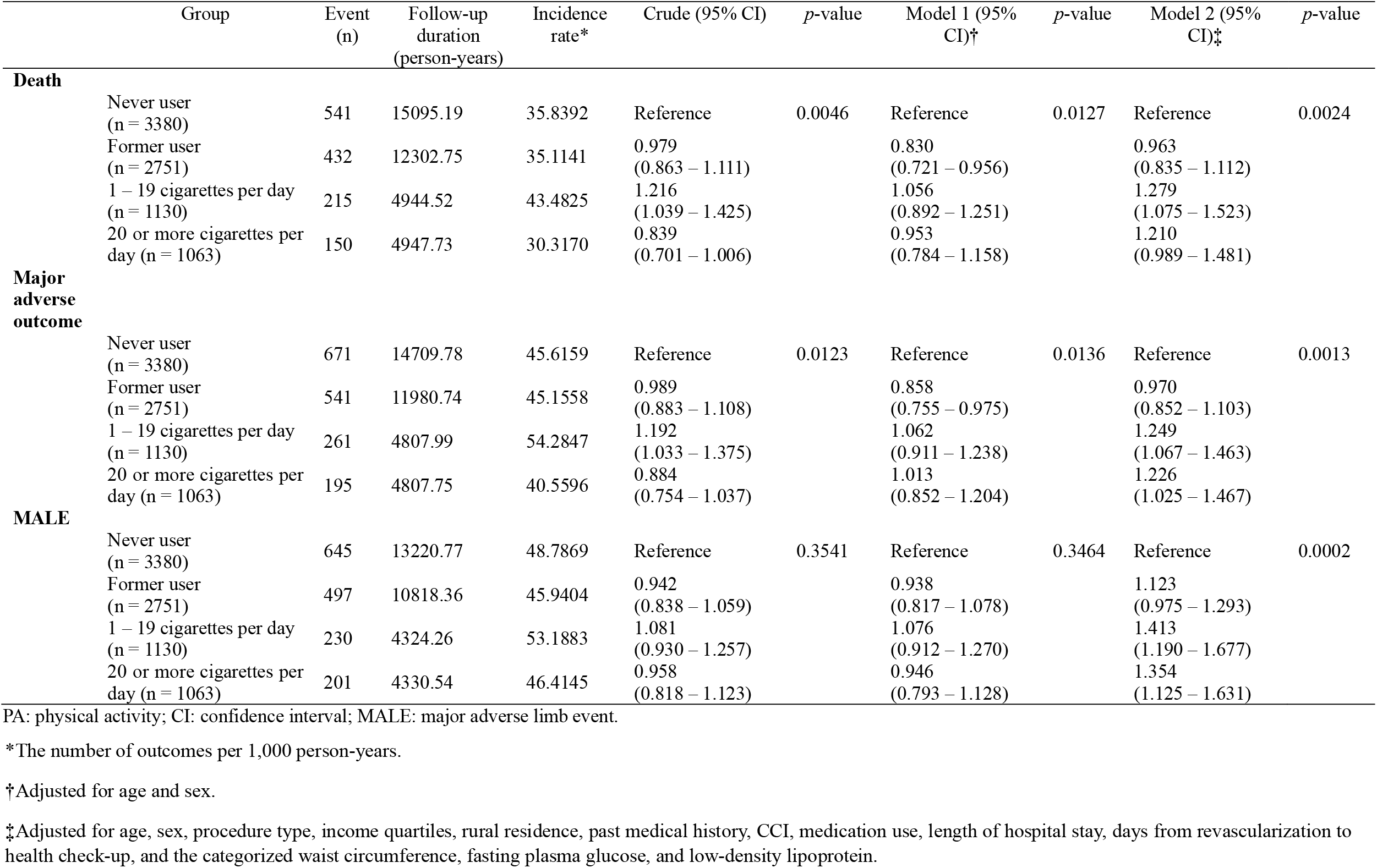
Hazard ratios for outcomes by tobacco use (further classification)

|  | Group | Event<br>(n) | Follow-up<br>duration<br>(person-years) | Incidence<br>rate* | Crude (95% CI) | *p*-value | Model 1 (95%<br>CI)<sup>†</sup> | *p*-value | Model 2 (95%<br>CI)<sup>‡</sup> | *p*-value |
| --- | --- | --- | --- | --- | --- | --- | --- | --- | --- | --- |
| **Death** | Never user<br>(n = 3380) | 541 | 15095.19 | 35.8392 | Reference | 0.0046 | Reference | 0.0127 | Reference | 0.0024 |
|  | Former user<br>(n = 2751) | 432 | 12302.75 | 35.1141 | 0.979<br>(0.863 – 1.111) |  | 0.830<br>(0.721 – 0.956) |  | 0.963<br>(0.835 – 1.112) |  |
|  | 1 – 19 cigarettes per day<br>(n = 1130) | 215 | 4944.52 | 43.4825 | 1.216<br>(1.039 – 1.425) |  | 1.056<br>(0.892 – 1.251) |  | 1.279<br>(1.075 – 1.523) |  |
|  | 20 or more cigarettes per<br>day (n = 1063) | 150 | 4947.73 | 30.3170 | 0.839<br>(0.701 – 1.006) |  | 0.953<br>(0.784 – 1.158) |  | 1.210<br>(0.989 – 1.481) |  |
| **Major**<br>**adverse**<br>**outcome** | Never user<br>(n = 3380) | 671 | 14709.78 | 45.6159 | Reference | 0.0123 | Reference | 0.0136 | Reference | 0.0013 |
|  | Former user<br>(n = 2751) | 541 | 11980.74 | 45.1558 | 0.989<br>(0.883 – 1.108) |  | 0.858<br>(0.755 – 0.975) |  | 0.970<br>(0.852 – 1.103) |  |
|  | 1 – 19 cigarettes per day<br>(n = 1130) | 261 | 4807.99 | 54.2847 | 1.192<br>(1.033 – 1.375) |  | 1.062<br>(0.911 – 1.238) |  | 1.249<br>(1.067 – 1.463) |  |
|  | 20 or more cigarettes per<br>day (n = 1063) | 195 | 4807.75 | 40.5596 | 0.884<br>(0.754 – 1.037) |  | 1.013<br>(0.852 – 1.204) |  | 1.226<br>(1.025 – 1.467) |  |
| **MALE** | Never user<br>(n = 3380) | 645 | 13220.77 | 48.7869 | Reference | 0.3541 | Reference | 0.3464 | Reference | 0.0002 |
|  | Former user<br>(n = 2751) | 497 | 10818.36 | 45.9404 | 0.942<br>(0.838 – 1.059) |  | 0.938<br>(0.817 – 1.078) |  | 1.123<br>(0.975 – 1.293) |  |
|  | 1 – 19 cigarettes per day<br>(n = 1130) | 230 | 4324.26 | 53.1883 | 1.081<br>(0.930 – 1.257) |  | 1.076<br>(0.912 – 1.270) |  | 1.413<br>(1.190 – 1.677) |  |
|  | 20 or more cigarettes per<br>day (n = 1063) | 201 | 4330.54 | 46.4145 | 0.958<br>(0.818 – 1.123) |  | 0.946<br>(0.793 – 1.128) |  | 1.354<br>(1.125 – 1.631) |  |

PA: physical activity; CI: confidence interval; MALE: major adverse limb event.
\*The number of outcomes per 1,000 person-years.
†Adjusted for age and sex.
‡Adjusted for age, sex, procedure type, income quartiles, rural residence, past medical history, CCI, medication use, length of hospital stay, days from revascularization to health check-up, and the categorized waist circumference, fasting plasma glucose, and low-density lipoprotein.

Finally, when patients were classified into four groups based on PA and tobacco status, active non-users showed significantly lower adjHRs than inactive tobacco users [death; 0.636 (0.531 – 0.761), major adverse outcome; 0.670 (0.569 – 0.789), MALE; 0.707 (0.590 – 0.846), **Table 5**]. The Kaplan-Meier analysis also showed a significant difference in clinical outcomes (**eFigure 8** in **Supplement**).

**Table 5.**
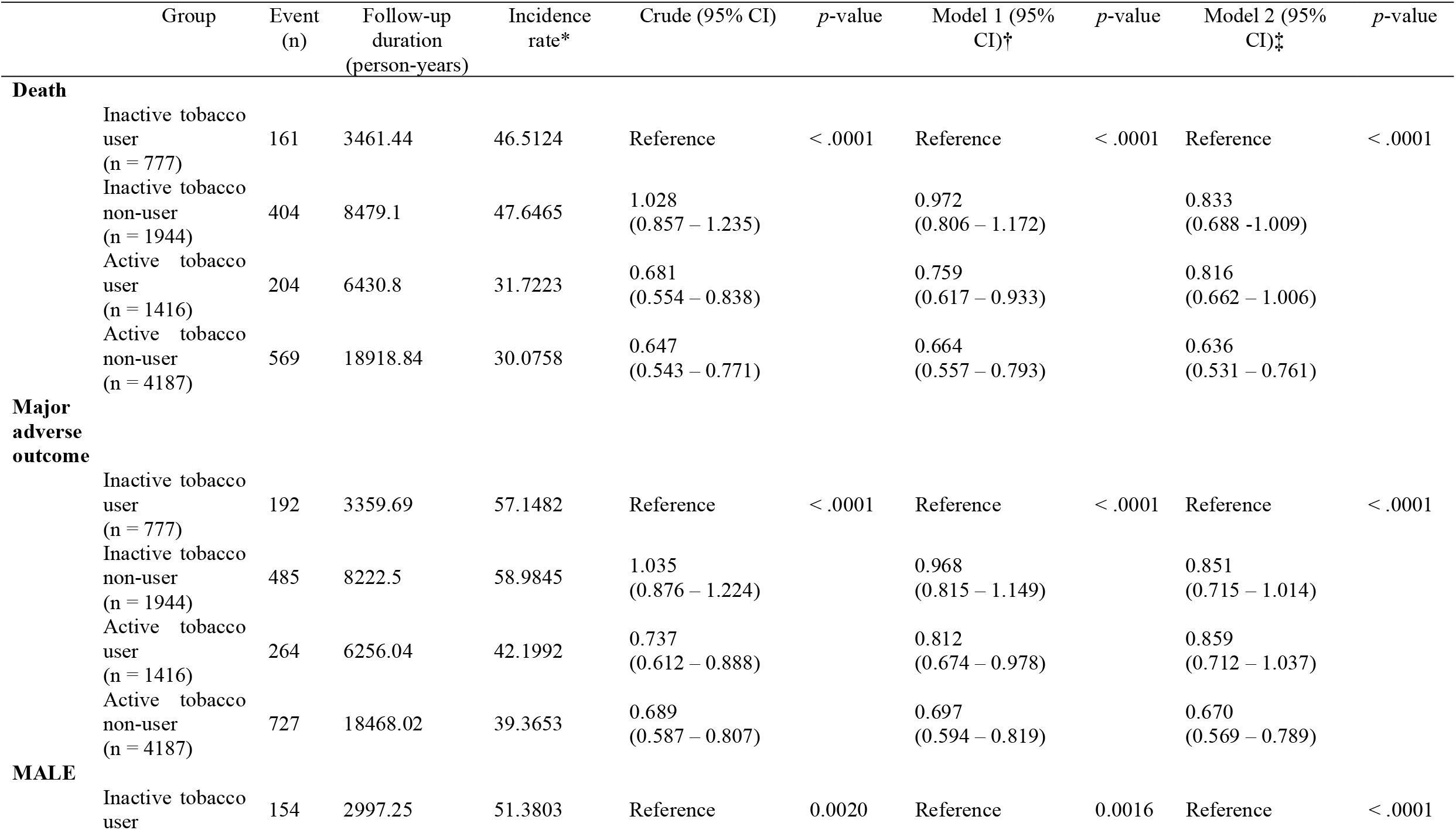
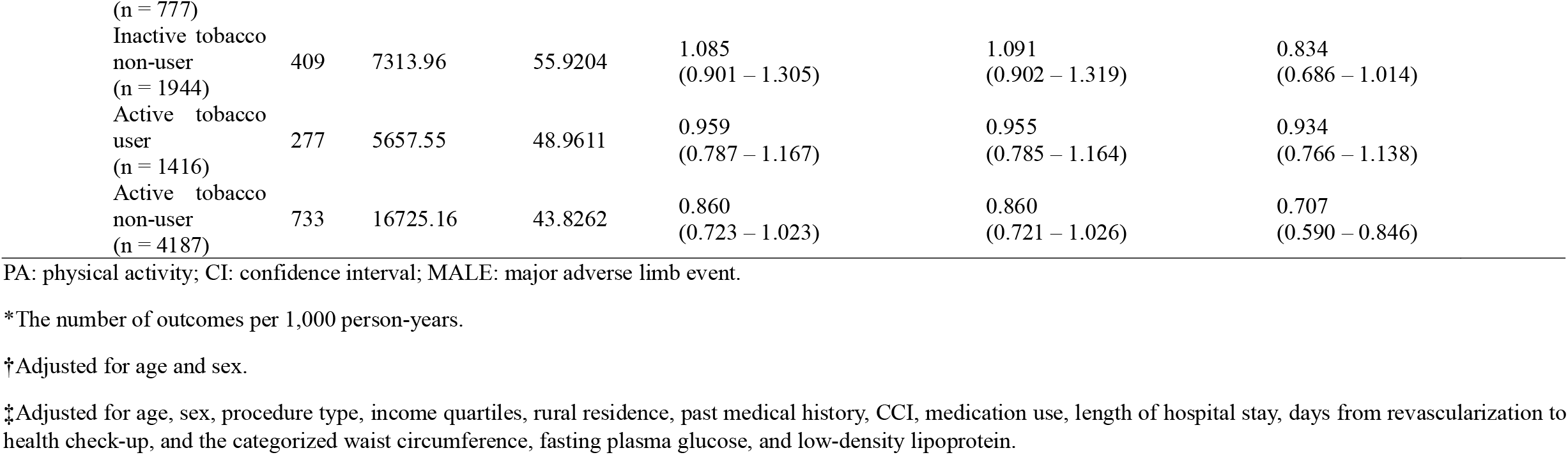
Hazard ratios for outcomes based on the combination of PA and tobacco use

## Discussion

The results of this population-based longitudinal cohort study in the Western Pacific region where PAD burden is highest revealed that modifiable risk factors were not well-addressed after receiving revascularization; approximately one-third of patients with PAD were physically inactive and a quarter were using tobacco. Moreover, unfortunately these common health-risk behaviours were closely related to adverse clinical outcomes such as all-cause mortality, morbidity, and MALE; Active patients had 14%–23% lower adverse clinical outcomes than inactive ones, and tobacco users had 26%–29% higher adverse clinical outcomes than non-users after adjusting for multiple confounders. While these findings are concordant with those of a previous study^33^, this has now been established in a large number of patients with PAD who received revascularization therapy, using nationwide, representative data, and in a broad range of clinical outcomes, such as mortality, recurrent revascularization, and amputation, important to patients, clinician-scientists, and payers alike.

A clear dose-response relationship with PA was observed, underlining the robustness of this association. This may support that patients with PAD need to increase PA after revascularization. However, barriers to PA are greater in patients with PAD than in those with other chronic conditions. Cavalcante et al. reported that low income and education, diabetes mellitus, and low ankle-brachial index were associated with low PA, and common barriers included lack of physical energy and lack of green space^34^. Hence, it would be beneficial for patients to be prescribed individually-tailored exercise programs (i.e., taking into consideration patient’s exercise history, current ability, preferences, and pain) and to be supported to improve adherence and progress in their activity, such as is offered in cardiac rehabilitation. Indeed, the benefit of exercise therapy is even greater when supervised by trained personnel^35^. Further, cardiac rehabilitation promotes other secondary prevention modifications such as tobacco cessation^36^. Given the high prevalence of PAD in Korea, the high degree of health-related behaviour and the degree of impact where they are modified, achieving greater cardiac rehabilitation use has the potential to significantly reduce morbidity and associated health care costs.

Indeed, in our study, tobacco users had approximately 30% higher rates of death, major adverse outcome, and MALE than non-users, while the amount of tobacco use did not affect clinical outcomes significantly. In addition, active tobacco users showed approximately 20% higher mortality and morbidity than active non-users. The harmful consequences of tobacco are because of its effects on the endothelium, platelets, and coagulation^37^. Accordingly, tobacco users have been shown to have more severe claudication, reduced exercise capacity, and poorer peripheral circulation^38^. However, tobacco users can achieve improvements in physical function after exercise rehabilitation, although users often have greater impairment than non-users at baseline^39^. Complete cessation is far more beneficial than only decreasing the amount of tobacco use as seen in a previous study^40^. Repeated cessation advice, nicotine replacement therapy, and medication such as bupropion should be offered^41^, as provided in cardiac rehabilitation. A tobacco product tax increase also may be beneficial at the public policy level^42^.

This study has several limitations. First, the diagnosis of PAD was based on claim code data. However, we used intervention codes instead of diagnostic codes to increase reliability. Given the relatively high cost for revascularization procedures and the fact that input of claim codes for these procedures is mandatory for reimbursement in Korean NHIS, missingness should be minimal. This approach has been used successfully in a previous study^11^. Second, selection bias may be present, considering that patients who did not attend a health check-up were excluded; non-attendance may have been due to death, as well as physical or cognitive disabilities. In addition, patients who experienced MI, stroke, recurrent revascularization, or amputation before the health check-up (i.e., a washout period) were excluded. This explains why our study cohort had a relatively lower annual mortality rate of about 3.7% compared to previous studies^6,43^. To rule out any potential bias related to this, the analyses were performed without the washout period; results were consistent (**eTable** in **Supplement)**. Third, there may be other confounding factors that we did not incorporate such as extent of the lesion and severity or classification of PAD; we tried to include as many confounders for mortality and morbidity as possible in the analysis. In terms of medication use, we extracted claim data about lipid-lowering agents instead of statins alone. However, a recent study in patients with PAD showed that almost all lipid-lowering agents taken were statins^44^. Fourth, PA was assessed via a self-administered questionnaire; hence, the degree of PA may have been over-stated; however, previous studies have established the psychometric properties of the self-administered questionnaire^26,45^. Finally, the effects of cardiac rehabilitation were not evaluated. The reimbursement of cardiac rehabilitation did not start until February 2017 in Korea; hence, claim codes for cardiac rehabilitation in our study cohort could not be identified from the NHIS database. Although MI is the most common indication for cardiac rehabilitation, the nationwide cardiac rehabilitation participation rate after MI is only around 1.5% in Korea^46^; hence, the cardiac rehabilitation participation rate in PAD would be so small as to have negligible impact on the findings.

## Conclusions

After receiving revascularization therapy for PAD, 32.7% patients were inactive and 26.4% used tobacco. Inactive patients and tobacco users showed significantly higher adjusted hazard ratios for adverse clinical outcomes. Therefore, patients with PAD should be referred and encouraged to attend cardiac rehabilitation to address these modifiable risk factors, and hence improve outcomes.

### One-sentence summary

Physical activity and tobacco use were significantly associated with mortality and morbidity in patients with peripheral arterial disease after revascularization.

## Supporting information

eFigure 1

eFigure 2

eFigure 3

eFigure 4

eFigure 5

eFigure 6

eFigure 7

eFigure 8

eTable 1

## Data Availability

The data are available through the Korean National Health Insurance Sharing Service. Researchers who wish to access the data should apply on the website (https://nhiss.nhis.or.kr/bd/ay/bdaya001iv.do) and request access to NHIS-2020-1-339.

## Acknowledgements

This study used National Health Information Database (NHIS-2020-1-339) made by NHIS. The authors would like to thank the Korean National Health Insurance Service for supporting data access and analysis.

## Author contributions

Cha S, Grace SL, Han K-D, and Kim W-S contributed to the conception of the work. Cha S, Han K-D, Kim B, and Kim W-S contributed to the acquisition and analysis of data and all authors contributed to the interpretation of data for the work. Cha S, Grace SL, and Kim W-S drafted the manuscript. All authors critically revised the manuscript. All authors gave final approval after reviewing all aspects of work ensuring integrity and accuracy.

## References

1. Fowkes FGR, Rudan D, Rudan I, et al. Comparison of global estimates of prevalence and risk factors for peripheral artery disease in 2000 and 2010: A systematic review and analysis. Lancet. 2013;382(9901):1329–1340.

2. Song P, Rudan D, Zhu Y, et al. Global, regional, and national prevalence and risk factors for peripheral artery disease in 2015: an updated systematic review and analysis. Lancet Glob Health. 2019;7(8):e1020–e1030.

3. Maksimovic M, Vlajinac H, Marinkovic J, Kocev N, Voskresenski T, Radak D. Health-related quality of life among patients with peripheral arterial disease. Angiology. 2014;65(6):501–506.

4. Abaraogu UO, Ezenwankwo EF, Dall PM, Seenan CA. Living a burdensome and demanding life: A qualitative systematic review of the patients experiences of peripheral arterial disease. PLoS One. 2018;13(11):e0207456.

5. Criqui MH, Langer RD, Fronek A, et al. Mortality over a Period of 10 Years in Patients with Peripheral Arterial Disease. N Engl J Med. 1992;326(6):381–386.

6. Caro J, Migliaccio-Walle K, Ishak KJ, Proskorovsky I. The morbidity and mortality following a diagnosis of peripheral arterial disease: Long term follow-up of a large database. BMC Cardiovasc Disord. 2005;5:14.

7. Davies MG. Criticial limb ischemia: epidemiology. Methodist Debakey Cardiovasc J. 2012;8(4):10–14.

8. Gerhard-Herman MD, Gornik HL, Barrett C, et al. 2016 AHA/ACC guideline on the management of patients with lower extremity peripheral artery disease: Executive Summary: A report of the American college of cardiology/American Heart Association task force on clinical practice guidelines. Circulation. 2017;135(12):e686–e725.

9. Aboyans V, Ricco JB, Bartelink Mlel, et al. 2017 ESC Guidelines on the Diagnosis and Treatment of Peripheral Arterial Diseases, in collaboration with the European Society for Vascular Surgery (ESVS). Eur Heart J. 2018;39(9):763–816.

10. Goodney PP, Beck AW, Nagle J, Welch HG, Zwolak RM. National trends in lower extremity bypass surgery, endovascular interventions, and major amputations. J Vasc Surg. 2009;50(1):54–60.

11. Park YY, Joh JH, Han SA, et al. National trends for the treatment of peripheral arterial disease in Korea between 2004 and 2013. Ann Surg Treat Res. 2015;89(6):319–324.

12. Lane R, Harwood A, Watson L, Leng GC. Exercise for intermittent claudication. Cochrane Database Syst Rev. 2017;12(12):CD000990.

13. Kruidenier LM, Nicola SP, Rouwet EV, Peters RJ, Prins MH, Teijink JAW. Additional supervised exercise therapy after a percutaneous vascular intervention for peripheral arterial disease: A randomized clinical trial. J Vasc Interv Radiol. 2011;22(7):961–968.

14. Saratzis A, Paraskevopoulos I, Patel S, et al. Supervised Exercise Therapy and Revascularization for Intermittent Claudication. JACC Cardiovasc Interv. 2019;12(12):1125–1136.

15. Frank U, Nikol S, Belch J, et al. ESVM Guideline on peripheral arterial disease. Vasa. 2019;48:1–79.

16. Smith SC, Benjamin EJ, Bonow RO, et al. AHA/ACCF secondary prevention and risk reduction therapy for patients with coronary and other atherosclerotic vascular disease: 2011 update: A guideline from the American Heart Association and American College of Cardiology Foundation. Circulation. 2011;124(22):2458–2473.

17. McDermott MMG, Liu K, Greenland P, et al. Functional decline in peripheral arterial disease: Associations with the ankle brachial index and leg symptoms. JAMA. 2004;292(4):453–461.

18. McDermott MMG, Liu K, O’Brien E, et al. Measuring physical activity in peripheral arterial disease: A comparison of two physical activity questionnaires with an accelerometer. Angiology. 2000;51(2):91– 100.

19. Lu L F Mackay D, Pell JP. Meta-analysis of the association between cigarette smoking and peripheral arterial disease. Heart. 2014;100(5):414–423.

20. Álvarez LR, Balibrea JM, Suriñach JM, et al. Smoking cessation and outcome in stable outpatients with coronary, cerebrovascular, or peripheral artery disease. Eur J Prev Cardiol. 2013;20(3):486–495.

21. Armstrong EJ, Wu J, Singh GD, et al. Smoking cessation is associated with decreased mortality and improved amputation-free survival among patients with symptomatic peripheral artery disease. J Vasc Surg. 2014;60(6):1565–1571.

22. Ng M, Freeman MK, Fleming TD, et al. Smoking prevalence and cigarette consumption in 187 countries, 1980-2012. JAMA. 2014;311(2):183–192.

23. Song SO, Jung CH, Song YD, et al. Background and data configuration process of a nationwide population-based study using the Korean national health insurance system. Diabetes Metab J. 2014;38(5):395–403.

24. Lee J, Lee JS, Park SH, Shin SA, Kim KW. Cohort profile: The national health insurance service-national sample cohort (NHIS-NSC), South Korea. Int J Epidemiol. 2017;46(2):e15.

25. National Health Insurance Service (NHIS), Health Insurance Review and Assessment Service (HIRA). 2017 National Health Insurance Statistical Yearbook. Wonju: National Health Insurance Service, Health Insurance Review & Assessment Service; 2017.

26. Smith BJ, Marshall AL, Huang N. Screening for physical activity in family practice: Evaluation of two brief assessment tools. Am J Prev Med. 2005;29(4):256–264.

27. Milton K, Bull FC, Bauman A. Reliability and validity testing of a single-item physical activity measure. Br J Sports Med. 2011;45(3):203–208.

28. Kim SH, Cha S, Kang S, Han K, Paik NJ, Kim WS. High prevalence of physical inactivity after heart valve surgery and its association with long-term mortality: A nationwide cohort study. Eur J Prev Cardiol. 2020;2047487320903877.

29. Jeong SW, Kim SH, Kang SH, et al. Mortality reduction with physical activity in patients with and without cardiovascular disease. Eur Heart J. 2019;40(43):3547–3555.

30. Agarwal DP. Cardioprotective effects of light-moderate consumption of alcohol: A review of putative mechanisms. Alcohol Alcohol. 2002;37(5):409–415.

31. World Health Organization. The Asia-Pacific perspective: redefining obesity and its treatment. 2000.

32. Quan H, Sundararajan V, Halfon P, et al. Coding algorithms for defining comorbidities in ICD-9-CM and ICD-10 administrative data. Med Care. 2005;43(11):1130–1139.

33. Garg PK, Tian L, Criqui MH, et al. Physical activity during daily life and mortality in patients with peripheral arterial disease. Circulation. 2006;114(3):242–248.

34. Cavalcante BR, Farah BQ, dos A Barbosa JP, et al. Are the barriers for physical activity practice equal for all peripheral artery disease patients? Arch Phys Med Rehabil. 2015;96(2):248–252.

35. Hageman D, Fokkenrood HJ, Gommans LN, van den Houten MM, Teijink JA. Supervised exercise therapy versus home-based exercise therapy versus walking advice for intermittent claudication. Cochrane Database Syst Rev. 2018;4(4):CD005263.

36. Jansen SCP, Hoorweg BB, Hoeks SE, et al. A systematic review and meta-analysis of the effects of supervised exercise therapy on modifiable cardiovascular risk factors in intermittent claudication. J Vasc Surg. 2019;69(4):1293-1308.e2.

37. Davis JW, Shelton L, Eigenberg DA, Hignite CE, Watanabe IS. Effects of tobacco and non-tobacco cigarette smoking on endothelium and platelets. Clin Pharmacol Ther. 1985;37(5):529–533.

38. Gardner AW. The effect of cigarette smoking on exercise capacity in patients with intermittent claudication. Vasc Med. 1996;1(3):181–186.

39. Gardner AW, Killewich LA, Montgomery PS, Katzel LI. Response to exercise rehabilitation in smoking and nonsmoking patients with intermittent claudication. J Vasc Surg. 2004;39(3):531–538.

40. Inoue-Choi M, Christensen CH, Rostron BL, et al. Dose-Response Association of Low-Intensity and Nondaily Smoking With Mortality in the United States. JAMA Netw Open. 2020;3(6):e206436

41. Hobbs SD, Bradbury AW. Smoking cessation strategies in patients with peripheral arterial disease: An evidence-based approach. Eur J Vasc Endovasc Surg. 2003;26(4):341–347.

42. Levin SR, Hawkins SS, Farber A, et al. Siracuse JJ. Association of State Tobacco Control Policies on Active Smoking at the Time of Intervention for Intermittent Claudication. J Vasc Surg. 2020;S0741-5214(20)32271-0.

43. Sigvant B, Hasvold P, Kragsterman B, et al. Cardiovascular outcomes in patients with peripheral arterial disease as an initial or subsequent manifestation of atherosclerotic disease: Results from a Swedish nationwide study. J Vasc Surg. 2017;66(2):507-514.e1.

44. Choi SY, Yang BR, Kang HJ, Park KS, Kim HS. Contemporary use of lipid-lowering therapy for secondary prevention in Korean patients with atherosclerotic cardiovascular diseases. Korean J Intern Med. 2020;35(3):593–604.

45. Washburn RA, Heath GW, Jackson AW. Reliability and validity issues concerning large-scale surveillance of physical activity. Res Q Exerc Sport. 2000;71:S104–S113.

46. Kim SH, Ro JS, Kim Y, Leigh JH, Kim WS. Underutilization of hospital-based cardiac rehabilitation after acute myocardial infarction in Korea. J Korean Med Sci. 2020;35(30):e262.

