## Supplementary figures and images for "Effect of physical inactivity and tobacco use on mortality and morbidity in revascularized patients with peripheral arterial disease: A nationwide cohort study"

### eFigure 1

**eFigure 1.** Study design.

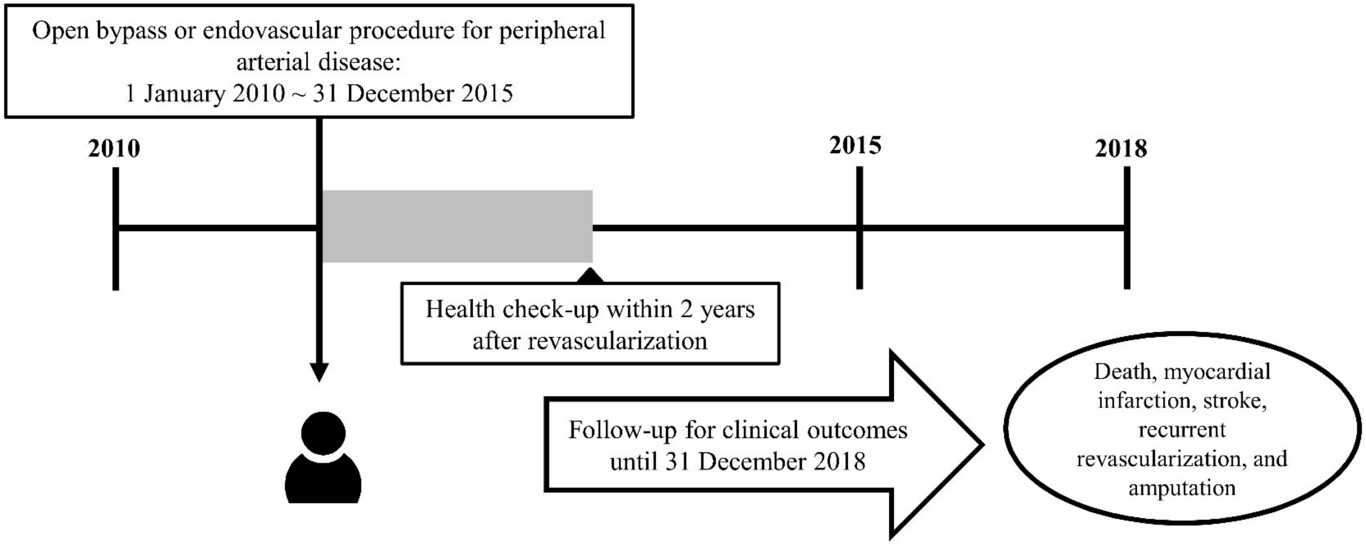

### eFigure 2

**eFigure 2.** Patient flow.

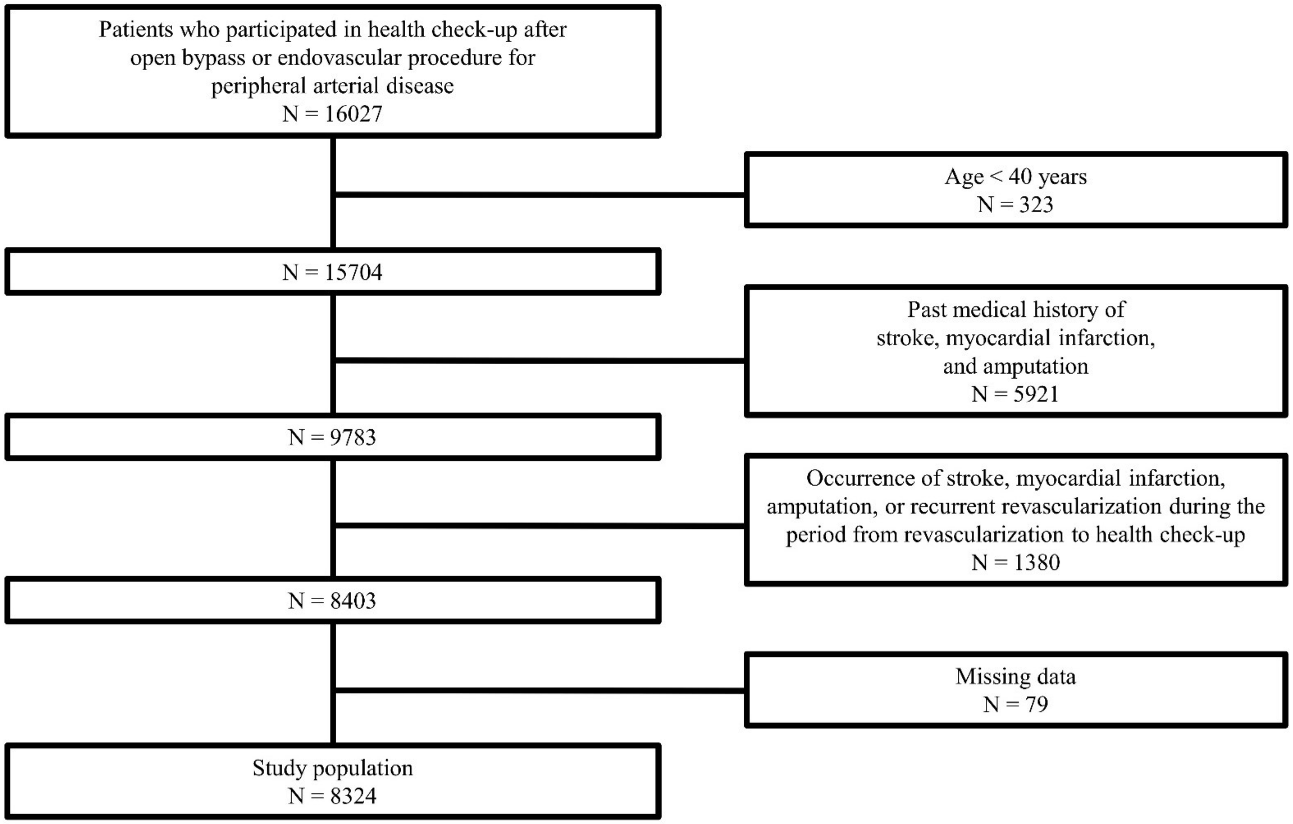
