## Supplementary material for "Effect of physical inactivity and tobacco use on mortality and morbidity in revascularized patients with peripheral arterial disease: A nationwide cohort study": eFigure 3

**eFigure 3.** Physical activity questionnaire.

Please read the following sentences and check the answer which corresponds to your physical activity status of the past week.

1. During the past week, how many days did you engage, for more than 20 minutes, in vigorous physical activities which made you notably more short of breath than usual? (e.g. running, aerobics, fast bicycling or mountain climbing)

   ☐0 ☐1 ☐2 ☐3 ☐4 ☐5 ☐6 ☐7

2. During the past week, how many days did you engage, for more than 30 minutes, in moderate physical activities which made you a little more short of breath than usual? (e.g. fast walking, doubles tennis or bicycling at a regular pace)

   ☐0 ☐1 ☐2 ☐3 ☐4 ☐5 ☐6 ☐7

3. During the past week, how many days did you walk at least 10 minutes at a time for a total of more than 30 minutes a day? (e.g. light exercise, including walking to and from work or leisure time walking)
   Exclude physical activities related to questions 1 and 2.

   ☐0 ☐1 ☐2 ☐3 ☐4 ☐5 ☐6 ☐7
