## Supplementary material for "Effect of physical inactivity and tobacco use on mortality and morbidity in revascularized patients with peripheral arterial disease: A nationwide cohort study": eFigure 4

**eFigure 4.** Physical activity categorization.

**Active**

1) At least 3 days of vigorous PA
2) At least 5 days of moderate PA
3) 4 days of moderate and 1 to 2 days of vigorous PA
4) 3 days of moderate and 2 days of vigorous PA
5) At least 5 days of walking
6) 4 days of walking and 1 to 4 days of moderate or vigorous PA
7) 3 days of walking and 2 to 4 days of moderate or vigorous PA
8) 2 days of walking and 3 to 4 days of moderate or vigorous PA
9) 1 days of walking and 4 days of moderate or vigorous PA

**Inactive**

Others
