## Supplementary material for "Effect of physical inactivity and tobacco use on mortality and morbidity in revascularized patients with peripheral arterial disease: A nationwide cohort study": eFigure 5

**eFigure 5.** Kaplan-Meier curves for PA and tobacco use.

(A) mortality, (B) major adverse outcome, and (C) MALE for PA, and (D) mortality, (E) major adverse outcome, and (F) MALE according for tobacco use.

PA: physical activity; MALE: major adverse limb event.

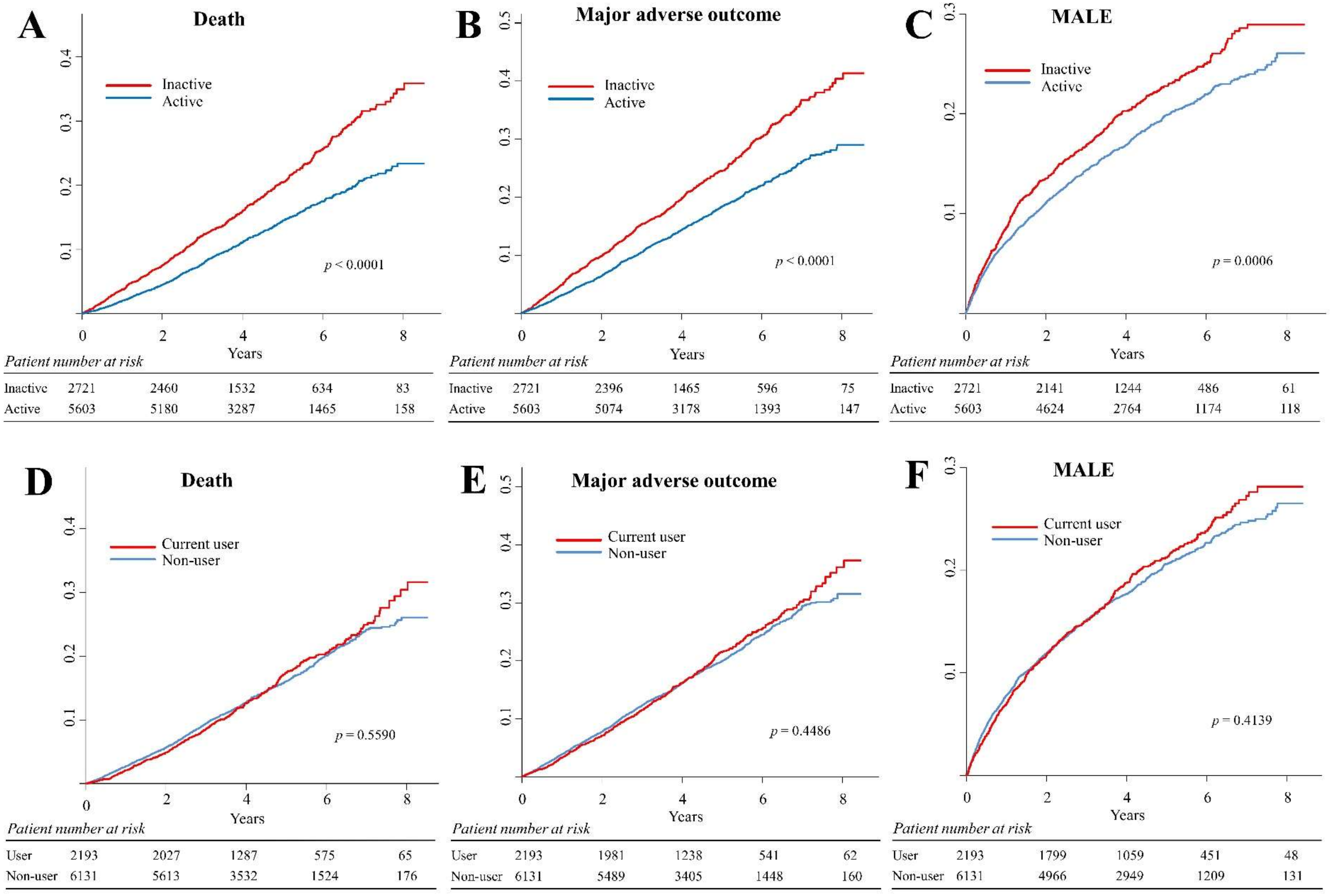
A
Death
Inactive
Active
p < 0.0001
Years
Patient number at risk
Inactive 2721 2460 1532 634 83
Active 5603 5180 3287 1465 158
B
Major adverse outcome
Inactive
Active
p < 0.0001
Years
Patient number at risk
Inactive 2721 2396 1465 596 75
Active 5603 5074 3178 1393 147
C
MALE
Inactive
Active
p = 0.0006
Years
Patient number at risk
Inactive 2721 2141 1244 486 61
Active 5603 4624 2764 1174 118
D
Death
Current user
Non-user
p = 0.5590
Years
Patient number at risk
User 2193 2027 1287 575 65
Non-user 6131 5613 3532 1524 176
E
Major adverse outcome
Current user
Non-user
p = 0.4486
Years
Patient number at risk
User 2193 1981 1238 541 62
Non-user 6131 5489 3405 1448 160
F
MALE
Current user
Non-user
p = 0.4139
Years
Patient number at risk
User 2193 1799 1059 451 48
Non-user 6131 4966 2949 1209 131
