## Supplementary material for "Effect of physical inactivity and tobacco use on mortality and morbidity in revascularized patients with peripheral arterial disease: A nationwide cohort study": eFigure 6

**eFigure 6.** Kaplan-Meier curves for PA-related energy expenditure.

(A) mortality, (B) major adverse outcome, and (C) MALE.

PA: physical activity; MALE: major adverse limb event.

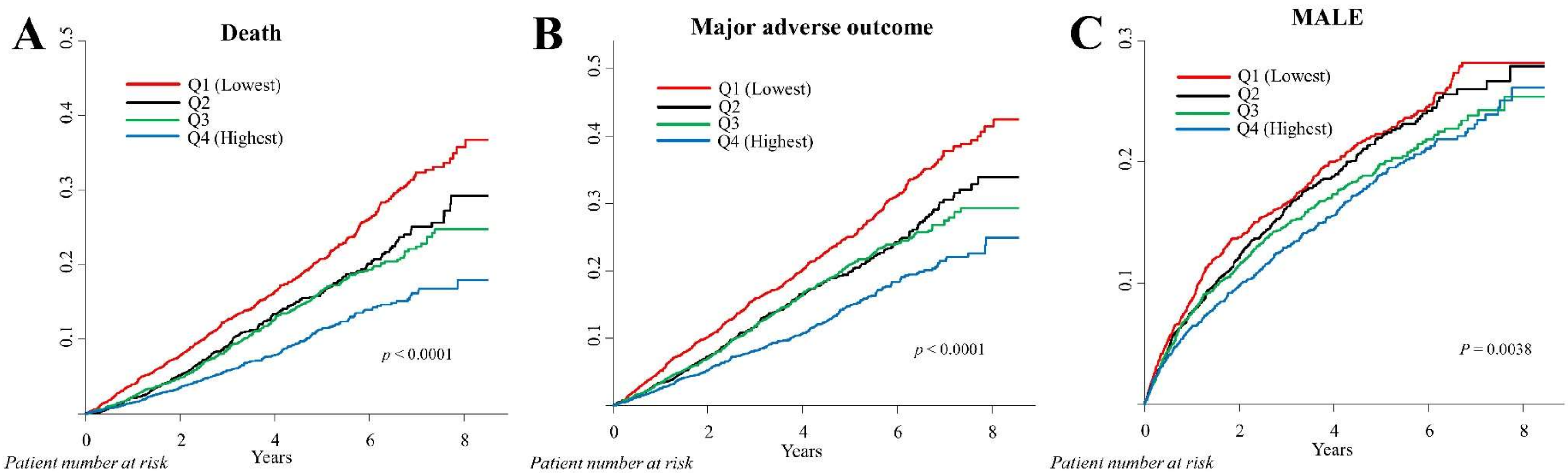


A — *Patient number at risk*

| | 0 | 2 | 4 | 6 | 8 |
|---|---|---|---|---|---|
| Q1 | 2441 | 2200 | 1373 | 569 | 74 |
| Q2 | 1696 | 1562 | 990 | 428 | 46 |
| Q3 | 2106 | 1936 | 1167 | 523 | 54 |
| Q4 | 2081 | 1942 | 1289 | 579 | 67 |

B — *Patient number at risk*

| | 0 | 2 | 4 | 6 | 8 |
|---|---|---|---|---|---|
| Q1 | 2441 | 2143 | 1312 | 534 | 67 |
| Q2 | 1696 | 1528 | 954 | 407 | 44 |
| Q3 | 2106 | 1890 | 1123 | 496 | 51 |
| Q4 | 2081 | 1909 | 1254 | 552 | 60 |

C — *Patient number at risk*

| | 0 | 2 | 4 | 6 | 8 |
|---|---|---|---|---|---|
| Q1 | 2441 | 1910 | 1122 | 440 | 54 |
| Q2 | 1696 | 1381 | 817 | 334 | 38 |
| Q3 | 2106 | 1718 | 973 | 415 | 37 |
| Q4 | 2081 | 1756 | 1096 | 471 | 50 |
