## Supplementary material for "Effect of physical inactivity and tobacco use on mortality and morbidity in revascularized patients with peripheral arterial disease: A nationwide cohort study": eFigure 7

**eFigure 7.** Kaplan-Meier curves for tobacco use (further classification).

(A) mortality, (B) major adverse outcome, and (C) MALE.

MALE: major adverse limb event.

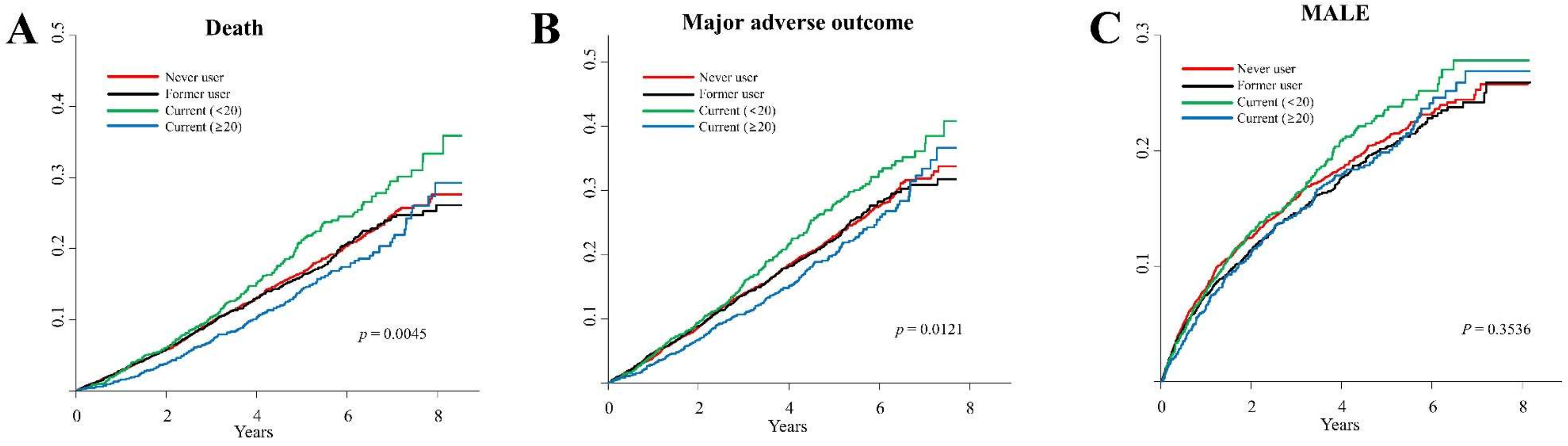


*Patient number at risk* (A)

| | | | | | |
|---|---|---|---|---|---|
| Never user | 3380 | 3097 | 1937 | 837 | 91 |
| Former user | 2751 | 2516 | 1595 | 687 | 85 |
| Current (<20) | 1130 | 1027 | 621 | 277 | 31 |
| Current (≥20) | 1063 | 1000 | 666 | 298 | 34 |

*Patient number at risk* (B)

| | | | | | |
|---|---|---|---|---|---|
| Never user | 3380 | 3033 | 1870 | 794 | 82 |
| Former user | 2751 | 2456 | 1535 | 654 | 78 |
| Current (<20) | 1130 | 805 | 597 | 259 | 31 |
| Current (≥20) | 1063 | 811 | 641 | 282 | 31 |

*Patient number at risk* (C)

| | | | | | |
|---|---|---|---|---|---|
| Never user | 3380 | 2725 | 1615 | 664 | 66 |
| Former user | 2751 | 2241 | 1334 | 545 | 65 |
| Current (<20) | 1130 | 905 | 513 | 218 | 25 |
| Current (≥20) | 1063 | 894 | 546 | 233 | 23 |
