## Supplementary material for "Effect of physical inactivity and tobacco use on mortality and morbidity in revascularized patients with peripheral arterial disease: A nationwide cohort study": eFigure 8

**eFigure 8.** Kaplan-Meier curves based on the combination of PA and tobacco use.

(A) mortality, (B) major adverse outcome, and (C) MALE.

PA: physical activity; MALE: major adverse limb event.

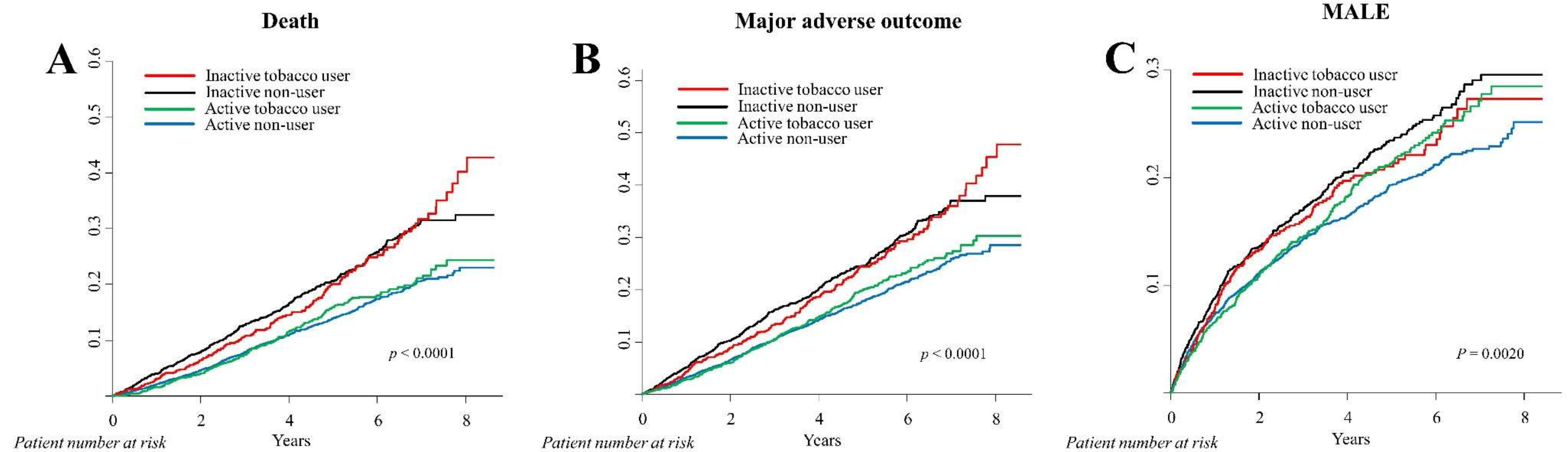


*Patient number at risk*

| | 0 | 2 | 4 | 6 | 8 |
|---|---|---|---|---|---|
| Inactive user | 777 | 710 | 446 | 191 | 26 |
| Inactive non-user | 1944 | 1750 | 1086 | 443 | 57 |
| Active user | 1416 | 1317 | 841 | 384 | 39 |
| Active non-user | 4187 | 3863 | 2446 | 1081 | 119 |

*Patient number at risk*

| | 0 | 2 | 4 | 6 | 8 |
|---|---|---|---|---|---|
| Inactive user | 777 | 692 | 425 | 181 | 24 |
| Inactive non-user | 1944 | 1704 | 1040 | 415 | 51 |
| Active user | 1416 | 1289 | 813 | 360 | 38 |
| Active non-user | 4187 | 3785 | 2365 | 1033 | 109 |

*Patient number at risk*

| | 0 | 2 | 4 | 6 | 8 |
|---|---|---|---|---|---|
| Inactive user | 777 | 619 | 363 | 149 | 19 |
| Inactive non-user | 1944 | 1522 | 881 | 337 | 42 |
| Active user | 1416 | 1180 | 696 | 302 | 29 |
| Active non-user | 4187 | 3444 | 2068 | 872 | 89 |
