## Supplementary material for "Effect of physical inactivity and tobacco use on mortality and morbidity in revascularized patients with peripheral arterial disease: A nationwide cohort study": eTable 1

**eTable 1.** Hazard ratios for outcomes according to PA and tobacco use without a washout period

| | Group | Event (n) | Follow-up duration (person-years) | Incidence rate* | Crude (95% CI) | *p*-value | Model 1 (95% CI)† | *p*-value | Model 2 (95% CI)‡ | *p*-value |
|---|---|---|---|---|---|---|---|---|---|---|
| **Death** | | | | | | | | | | |
| PA | Active (n = 6458) | 953 | 28729.04 | 33.1720 | 0.652 (0.592 – 0.719) | < .0001 | 0.701 (0.636 – 0.773) | < .0001 | 0.762 (0.690 – 0.841) | < .0001 |
| | Inactive (n = 3228) | 707 | 13930.52 | 50.7519 | Reference | | Reference | | Reference | |
| Tobacco use | Current user (n = 2465) | 421 | 11046.17 | 38.1128 | 0.969 (0.868 – 1.082) | 0.5779 | 1.051 (0.937 – 1.178) | 0.3978 | 1.209 (1.073 – 1.361) | 0.0017 |
| | Non-user (n = 7221) | 1239 | 31613.39 | 39.1922 | Reference | | Reference | | Reference | |
| **Major adverse outcome** | | | | | | | | | | |
| PA | Active (n = 6458) | 1201 | 27988.43 | 42.9106 | 0.690 (0.631 – 0.753) | < .0001 | 0.736 (0.674 – 0.804) | < .0001 | 0.791 (0.723 – 0.865) | < .0001 |
| | Inactive (n = 3228) | 838 | 13493.99 | 62.1017 | Reference | | Reference | | Reference | |
| Tobacco use | Current user (n = 2465) | 526 | 10709.95 | 49.1132 | 0.997 (0.902 – 1.100) | 0.9453 | 1.081 (0.975 – 1.198) | 0.1402 | 1.217 (1.094 – 1.354) | 0.0003 |
| | Non-user (n = 7221) | 1513 | 30772.48 | 49.1673 | Reference | | Reference | | Reference | |
| **MALE** | | | | | | | | | | |
| PA | Active (n = 6458) | 1392 | 24665.70 | 56.4347 | 0.830 (0.761 – 0.906) | < .0001 | 0.829 (0.760 – 0.905) | < .0001 | 0.881 (0.807 – 0.962) | 0.0049 |
| | Inactive (n = 3228) | 802 | 11625.27 | 68.9877 | Reference | | Reference | | Reference | |
| Tobacco use | Current user (n = 2465) | 546 | 9479.29 | 57.5992 | 0.941 (0.854 – 1.037) | 0.2196 | 0.966 (0.873 – 1.069) | 0.5006 | 1.223 (1.100 – 1.359) | 0.0002 |
| | Non-user (n = 7221) | 1648 | 26811.67 | 61.4658 | Reference | | Reference | | Reference | |

PA: physical activity; CI: confidence interval; MALE: major adverse limb event.

*The number of outcomes per 1,000 person-years.

†Adjusted for age and sex.

‡Adjusted for age, sex, procedure type, income quartiles, rural residence, past medical history, CCI, medication use, length of hospital stay, days from revascularization to health check-up, and the categorized waist circumference, fasting plasma glucose, and low-density lipoprotein.
